# Long-Term Persistence of Spike Antibody and Predictive Modeling of Antibody Dynamics Following Infection with SARS-CoV-2

**DOI:** 10.1101/2020.11.20.20235697

**Authors:** Louis Grandjean, Anja Saso, Arturo Torres Ortiz, Tanya Lam, James Hatcher, Rosie Thistlethwayte, Mark Harris, Timothy Best, Marina Johnson, Helen Wagstaffe, Elizabeth Ralph, Annabelle Mai, Caroline Colijn, Judith Breuer, Matthew Buckland, Kimberly Gilmour, David Goldblatt, the Co-Stars Study Team

**Affiliations:** Department of Infection, Inflammation and Immunity, Great Ormond Street Institute of Child Health, University College London, 30 Guilford Street, London, UK WC1N 1EH; Department of Infectious Diseases, Great Ormond Street Hospital, Great Ormond Street, London, WC1N 3JH; Department of Tropical and Infectious diseases; LSHTM, Keppel St, Bloomsbury, London WC1E 7HT; MRC Gambia at LSHTM, PO Box 273, Fajara, The Gambia; Department of Medicine, Imperial College, Paddington, London, W2 1NY; Department of Microbiology, Great Ormond Street Hospital, Great Ormond Street, London, WC1N 3JH; Management, Great Ormond Street Hospital, Great Ormond Street, London, WC1N 3JH; Quality Improvement, Great Ormond Street Hospital, Great Ormond Street, London, WC1N 3JH; Clinical Immunology, Camelia Botnar Laboratories, Great Ormond Street Hospital, Great Ormond Street, London, WC1N 3JH; Department of Mathematics, Simon Fraser University, Vancouver, British Colombia, Canada BC V5A

**Keywords:** Immunity, serology, antibody, ELISA, kinetics, neutralization, SARS-CoV-2, COVID-19, virus, nucleoprotein, spike protein

## Abstract

**Background:** Antibodies to Severe Acute Respiratory Syndrome Coronavirus-2 (SARS-CoV-2) have been shown to neutralize the virus *in-vitro*. Similarly, animal challenge models suggest that neutralizing antibodies isolated from SARS-CoV-2 infected individuals prevent against disease upon re-exposure to the virus. Understanding the nature and duration of the antibody response following SARS-CoV-2 infection is therefore critically important.

**Methods:** Between April and October 2020 we undertook a prospective cohort study of 3555 healthcare workers in order to elucidate the duration and dynamics of antibody responses following infection with SARS-CoV-2. After a formal performance evaluation against 169 PCR confirmed cases and negative controls, the Meso-Scale Discovery assay was used to quantify in parallel, antibody titers to the SARS-CoV-2 nucleoprotein (N), spike (S) protein and the receptor-binding-domain (RBD) of the S-protein. All seropositive participants were followed up monthly for a maximum of 7 months; those participants that were symptomatic, with known dates of symptom-onset, seropositive by the MSD assay and who provided 2 or more monthly samples were included in the analysis. Survival analysis was used to determine the proportion of sero-reversion (switching from positive to negative) from the raw data. In order to predict long-term antibody dynamics, two hierarchical longitudinal Gamma models were implemented to provide predictions for the lower bound (continuous antibody decay to zero, “Gamma-decay”) and upper bound (decay-to-plateau due to long lived plasma cells, “Gamma-plateau”) long-term antibody titers.

**Results:** A total of 1163 samples were provided from 349 of 3555 recruited participants who were symptomatic, seropositive by the MSD assay, and were followed up with 2 or more monthly samples. At 200 days post symptom onset, 99% of participants had detectable S-antibody whereas only 75% of participants had detectable N-antibody. Even under our most pessimistic assumption of persistent negative exponential decay, the S-antibody was predicted to remain detectable in 95% of participants until 465 days [95% CI 370-575] after symptom onset. Under the Gamma-plateau model, the entire posterior distribution of S-antibody titers at plateau remained above the threshold for detection indefinitely. Surrogate neutralization assays demonstrated a strong positive correlation between antibody titers to the S-protein and blocking of the ACE-2 receptor *in-vitro* [R^2^=0.72, p<0.001]. By contrast, the N-antibody waned rapidly with a half-life of 60 days [95% CI 52-68].

**Discussion:** This study has demonstrated persistence of the spike antibody in 99% of participants at 200 days following SARS-CoV-2 symptoms and rapid decay of the nucleoprotein antibody. Diagnostic tests or studies that rely on the N-antibody as a measure of seroprevalence must be interpreted with caution. Our lowest bound prediction for duration of the spike antibody was 465 days and our upper bound predicted spike antibody to remain indefinitely in line with the long-term seropositivity reported for SARS-CoV infection. The long-term persistence of the S-antibody, together with the strong positive correlation between the S-antibody and viral surrogate neutralization *in-vitro*, has important implications for the duration of functional immunity following SARS-CoV-2 infection.

## Introduction

Since appearing as a cluster of pneumonia cases in December 2019 in Wuhan, China, Coronavirus disease (COVID-19) has rapidly spread worldwide^1^. As of October 26^th^, there have been 43,187,134 cases, resulting in over 1.1 million deaths and a global health crisis, with significant social, economic and public health implications^2^. COVID-19 is caused by severe acute respiratory syndrome coronavirus-2 (SARS-CoV-2), an enveloped RNA β-coronavirus^3^. Specific immunoglobulin (IgG) antibody responses to the SARS-CoV-2 trimeric spike (S) protein, nucleoprotein (N) protein and the receptor-binding domain (RBD) develop between 6-15 days following disease-onset^4^. The S-protein, which contains the RBD, binds to host cells via the angiotensin-converting-enzyme-2 (ACE-2) receptor, and membrane fusion occurs before viral entry^5,6^. The N-protein plays an important role in transcription enhancement and viral assembly^7^.

SARS-CoV-2-specific antibodies, particularly to the S- and RBD-antigens, have been shown to correlate with T-cell responses and viral neutralization *in vitro* as well as to protect against disease in animals, following passive transfer of convalescent serum or selected monoclonal antibodies^8–12^. It is unclear, however, whether re-infection can occur in humans who mount a humoral response following primary SARS-CoV-2 infection and achieve viral clearance. Recent case reports have emerged describing new respiratory samples positive for SARS-CoV-2 RNA after confirmed negativity, although these are few compared to the worldwide scale of infection, with several potential explanations proposed^13,14^. Furthermore, no recurrence of disease was reported in rhesus macaques or Syrian hamsters that were re-challenged in the presence of detectable endogenous antibodies (although protection against infection varied between studies).^9,15,16^. These findings highlight the importance of characterising humoral dynamics following SARS-CoV-2 infection.

SARS-CoV IgG and neutralizing antibodies have been shown to commonly persist up to 2-3 years post-infection, particularly in hospitalized patients,^17,18^ with recent reports demonstrating seropositivity as late as 12-17 years after infection^19,20^. Following severe disease caused by Middle Eastern Respiratory Syndrome (MERS), antibodies have been detected up to 34-months post-infection^21,22^. Existing longitudinal studies of SARS-CoV-2 are limited by inadequate modeling of antibody dynamics, short duration, low sampling density and frequency of longer-term follow-up^23–32^. Fitting Locally Estimated Scatterplot Smoothing (LOESS) or equivalent lines of best fit to the data^23–25,33^ also fails to provide a mathematical framework for evaluating long-term antibody responses.

In order to evaluate antibody kinetics and longevity following SARS-CoV-2 infection, we undertook the prospective Covid-19 Staff Testing of Antibody Responses Study (Co-STARS). Seropositive and symptomatic participants were followed up monthly with repeated antibody titer quantification. Detailed demographic, clinical and socioeconomic data were collected and mathematical models developed to characterize longitudinal humoral kinetics from initial antibody boosting to subsequent decay. To predict long-term antibody dynamics, we fitted two different models based on the gamma distribution: one which assumed persistent antibody decay^34^, and an alternate that allowed for an eventual plateau,^35,36^ to account for sustained antibody production by long-lived plasma cells (LLPCs).

## Results

### Participant Demographics

After providing informed consent, a total of 3555 individuals - all healthcare workers at Great Ormond Street Hospital - were enrolled in the study. Of this group, 349 were both symptomatic, seropositive by the MSD assay and provided 2 or more monthly samples for the primary outcome analysis of antibody dynamics. These 349 seropositive participants were followed up monthly for a maximum of 7 months and provided 1163 serial monthly serological samples. The median follow-up time per participant was 122 days (IQR 65 – 157 days) with a maximum follow up time of 262 days from symptom onset. The majority of participants 252/349 (72%) donated 3 or more samples with a maximum of 7 samples donated during follow up. Most seropositive participants were women (80%) with a mean age of 39 years representative of the underlying population structure of the hospital. The predominant symptoms reported were cough 225/349 (64%), myalgia 225/349 (64%), followed by ageusia and anosmia at 210/349 (60%) and 201/349 (58%) respectively.

### Factors Associated with Increased Peak Antibodies and Rapid Decay

Multivariate analysis demonstrated that fever, rigors, ageusia, anosmia, a previous medical condition, high BMI and Black Asian Minority Ethnic (BAME) backgrounds were all associated with higher peak spike protein antibody titers (Table 1). No variables were identified to be independently associated with the rate of antibody decay.

**Table 1.**
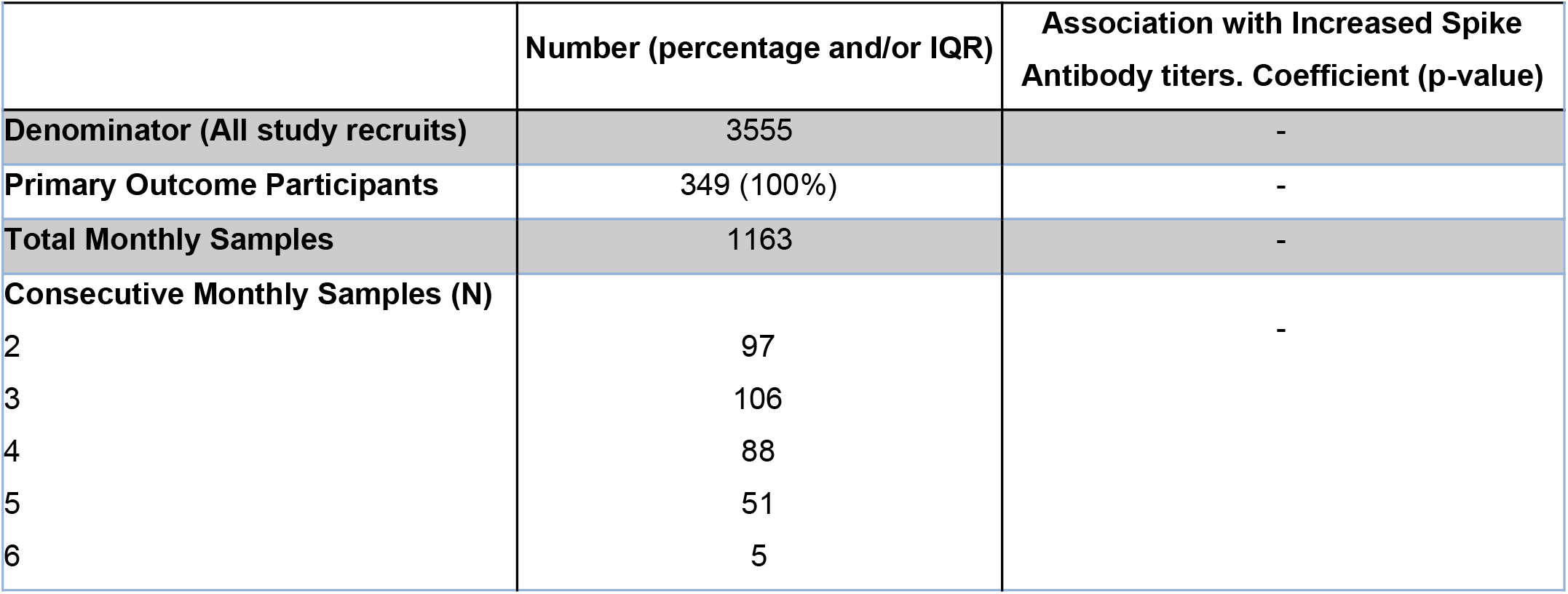
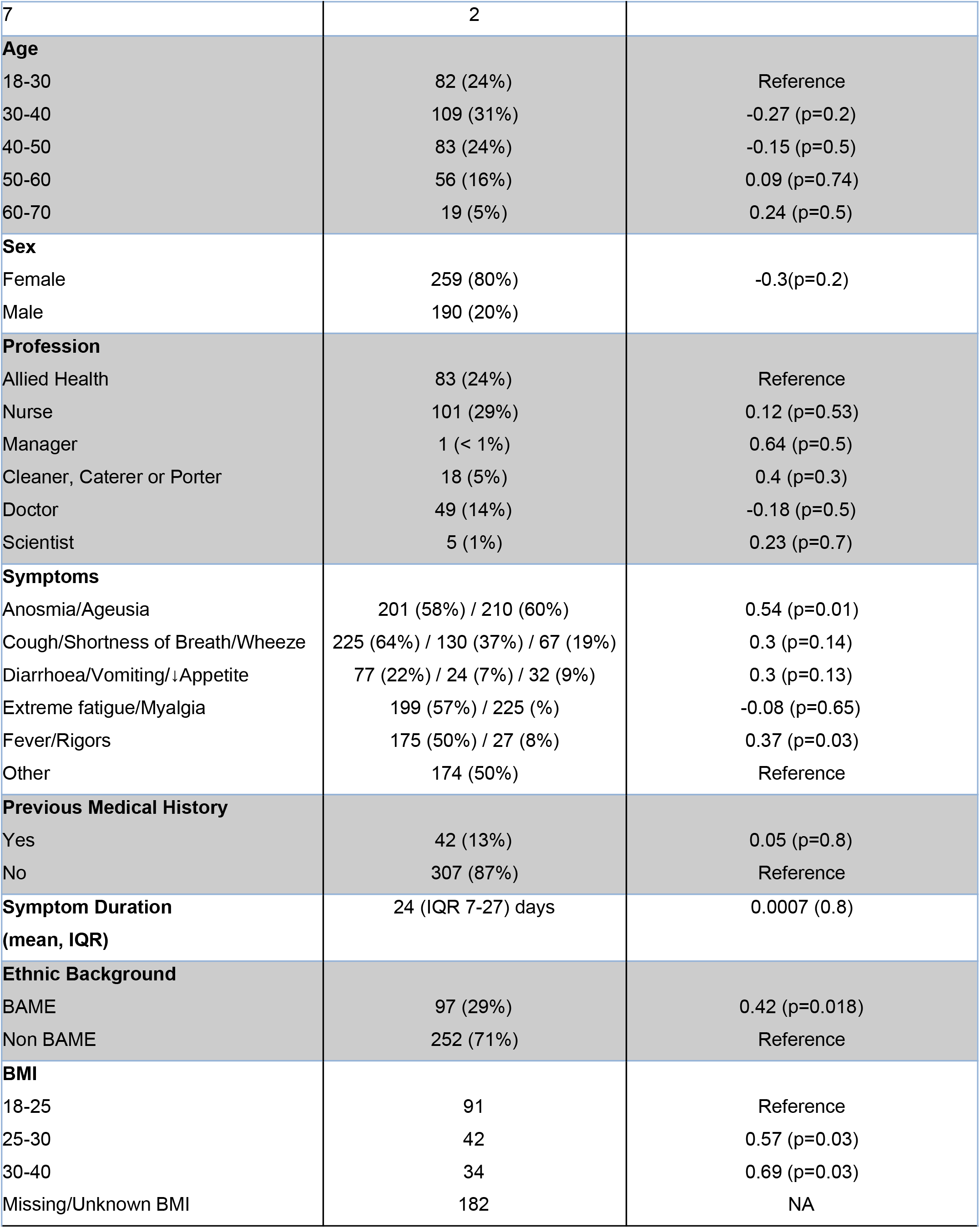
Demographic details of study participants and variables associated with high peak antibody titers.

### Observed Antibody Kinetics and Seroreversion

Serial monthly serological measurements from 349 participants who provided 2 or more samples following the onset of symptoms demonstrated a rapid rate of decay of the N-antibody relative to the S and RBD antibody (Figure 1). The spike antibody assay detected a total of 342/349 (98%) participants who were seropositive to any one of the S, RBD or N-antibodies. In comparison the RBD and N-assays detected 332/349 (95%) and 333/349 (95%) respectively. The sensitivity of the RBD and N-assays further declined with time relative to the S-antibody assay. At 200 days following the onset of symptoms, only 75% of 349 participants tested positive for N-antibody whereas 99% remained positive for S-antibody (Figure 2).

**Figure 1.**
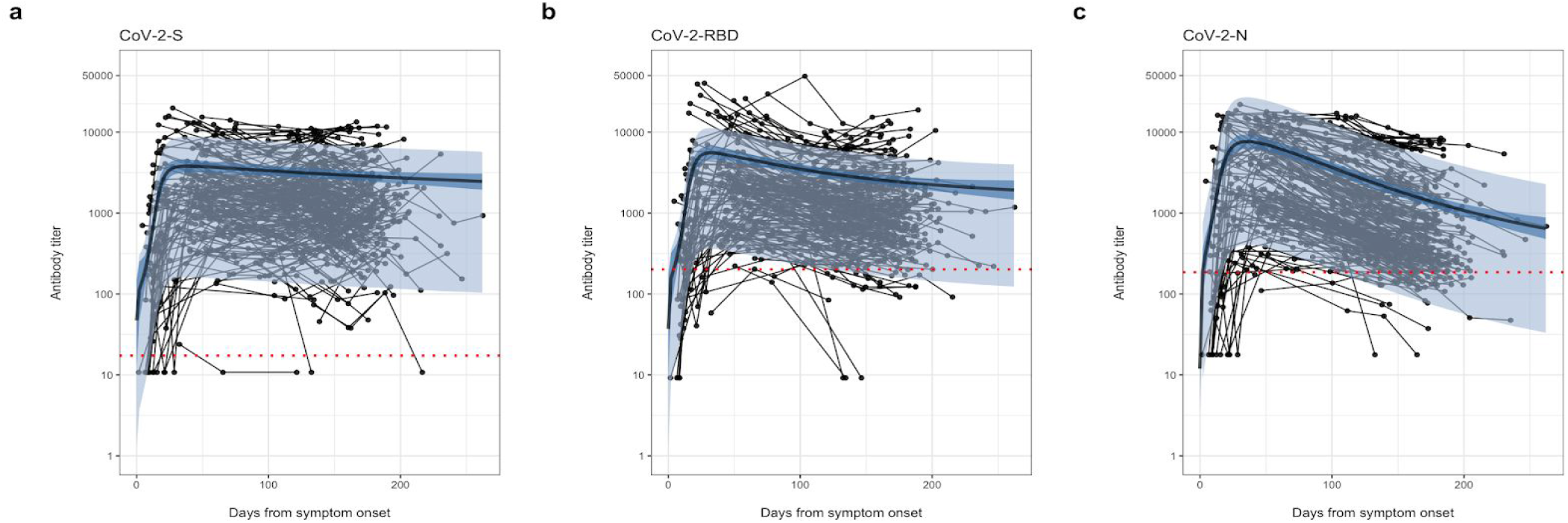
Serial monthly serological measurements from 349 participants up to 262 days following symptoms of SARS-CoV-2. Samples from the same participant are linked with a thin black line and the red-dotted line is shown to indicate seroreversion. The gamma-plateau model is superimposed to show antibody trajectory: The predicted antibody trajectory (black line) is the median of the posterior distribution of the best model fit and 95% CI with and without individual effects (light blue and dark blue shading respectively) **a)** The spike (S) protein **b)** RBD antibody and **c)** the nucleoprotein (N) antibody with a relatively steep rate of decay.

**Figure 2.**
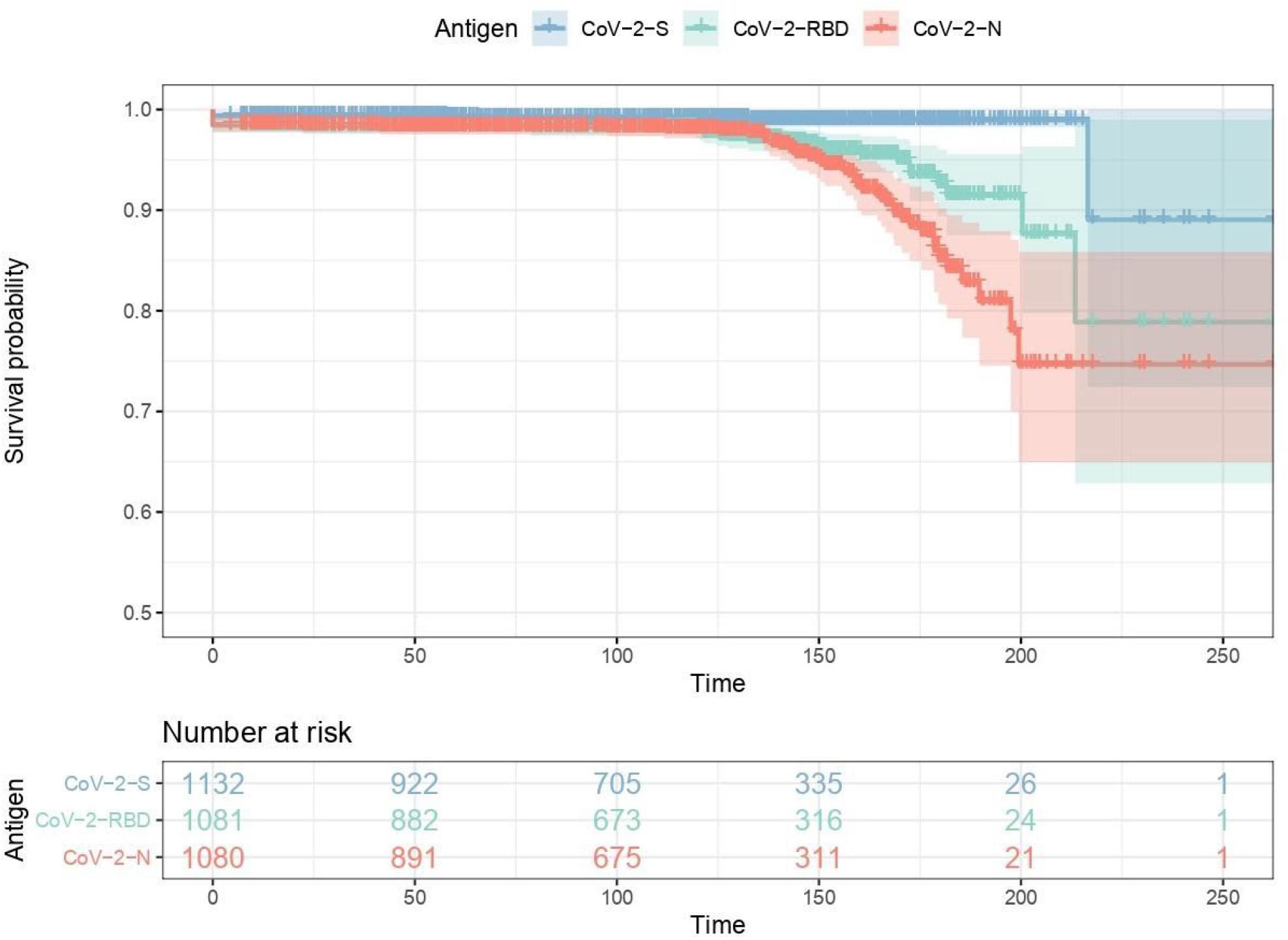
Parallel serological measurements of the Spike, RBD and Nucleoprotein antibodies from the start of symptoms. Repeated serological measurements to the spike (S, Blue), RBD (Green) and the nucleoprotein (N, Red) demonstrating the time to a negative test from all possible starting positive tests.

### Modeled Serological Reversion and Proportion of Positive Tests Over Time

Monte-Carlo Markov traces converged well for both the gamma distribution and decay-to-plateau curves demonstrating a stable model fit to the data (Supplementary Figure 1). The maximum R-hat for any parameter was 1.0035, while the minimum effective sample size (ESS) was 842.8 (Supplementary data, Table S1). Comparison of goodness of fit between models showed that for all antigens the decay-to-plateau model provided a better fit to the data than the gamma-decay model, although this difference was not statistically significant (Supplementary data, table S2). Even under the most pessimistic assumption of continuous gamma-decay we estimate that 95% of individuals following infection with SARS-CoV-2 will have measurable S-antibody until 465 days [95% CI 370-575] after the symptom start date. Under the gamma-plateau model S-antibody will remain detectable indefinitely, in line with recent reports on antibody kinetics following SARS-CoV-1 infection (Figure 3a)^20^. The most pessimistic gamma-decay model (lower bound) and most optimistic gamma-plateau model (upper bound) for each antibody are shown in Figure 3b. Under both models the N-antibody decayed to undetectable levels. Even after accounting for long-lived plasma cells under the gamma-plateau model, 75% of participants were predicted to have seroreverted N-antibody by 610 days [95% CI 420-530] whereas under the gamma-decay model 100% of participants had seroreverted N-antibody by 460 days [95% CI 420-530] following symptom onset.

**Figure 3:**
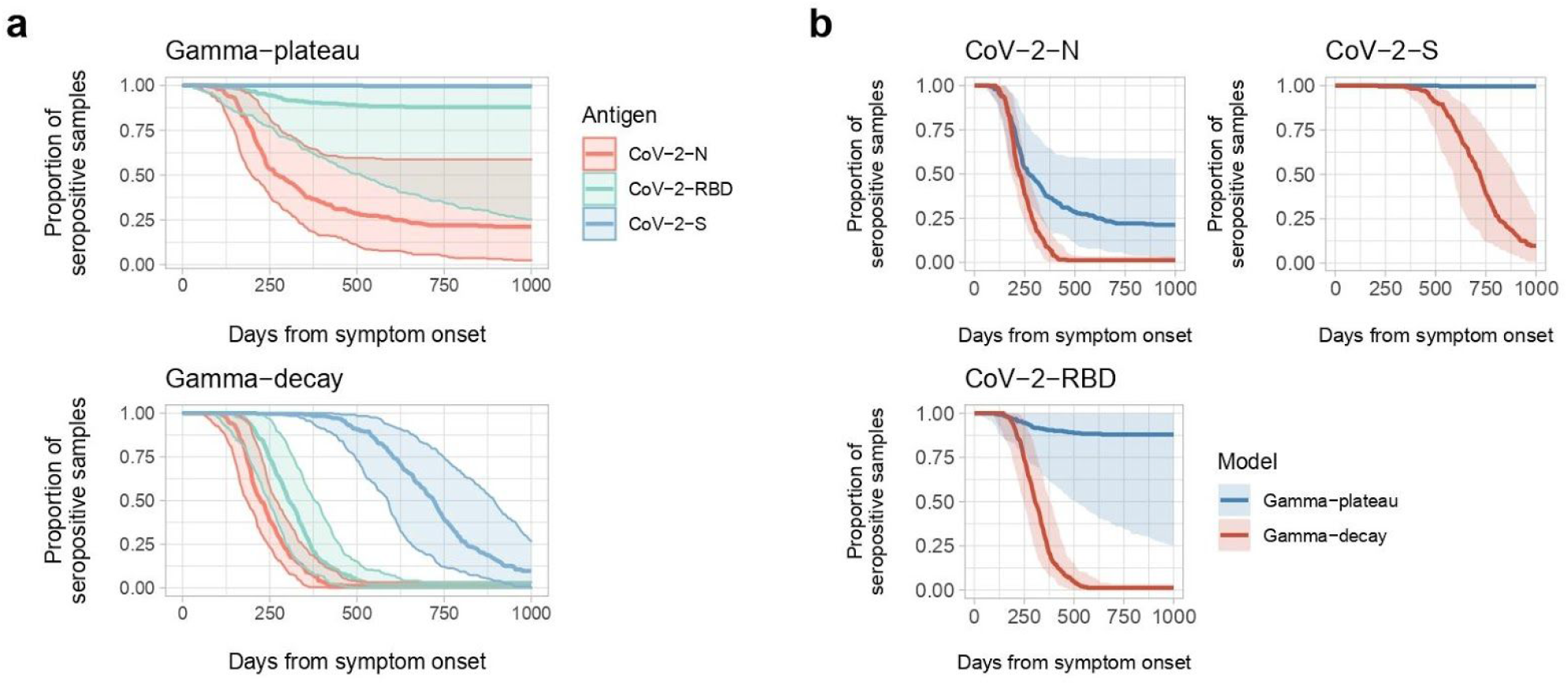
Modeled predicted time to seronegativity from symptom onset. Model based predictions of time to seronegativity. **a**, Comparison of the three tested antibodies against the S-protein (S, blue), receptor-binding domain (RBD, green) and nucleocapsid protein (N, red) for the gamma plateau model (top) and the gamma decay model (bottom). **b**, Differences between the two proposed models, the gamma plateau model (blue) and the gamma decay model (red) for the three tested antibodies (S; top right, RBD; bottom, and N; top left). Coloured lines represent the median estimates of the posterior density, while the shaded ribbons encompass the 95% confidence interval.

### Antibody Peak, Half-Life and Plateau

Measured weekly average titer data for each of the S-, RBD-, and N-antibodies demonstrated that peak antibody response to infection was itself a plateau/slowly increasing line. Antibody titers rapidly increased during the first 3 weeks with prolonged high titers reached and maintained between week 4 and week 10 after the onset of symptoms. The peak antibody response for the S-antibody, RBD, and N-antibody from both raw weekly average serial titer and modeled data occurred at 40 [95% CI 30-63] days, 31 days [95% CI 26-38], and 35 [95% CI 31-42] days respectively. This was supported by the both the gamma-decay and gamma-plateau models which provided a similar close fit to this early stage of the humoral response (Figure 4 a, b, c).

**Figure 4.**
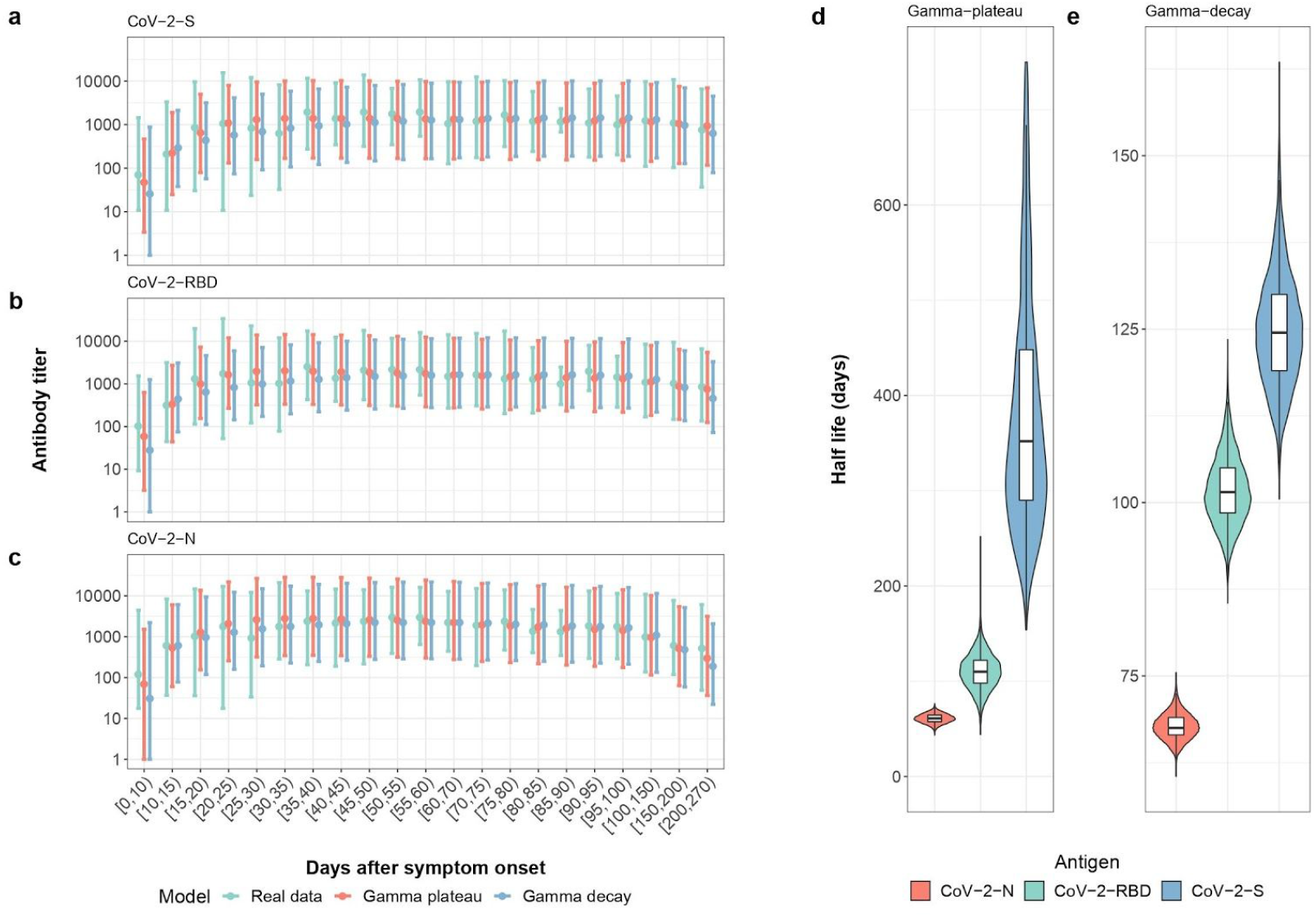
**a, b, c, d, e) Measured and Modeled Weekly Mean Antibody Titer**. Real data (green), gamma-plateau model (red), gamma-decay model (blue) for **a)** the spike antibody, **b)** the RBD antibody and **c)** the N-antibody. **Modeled half-lives of antibody decay for d)** The gamma-plateau model and **e)** gamma-decay model. Colors represent the three different antibodies tested: those for the spike protein (blue), nucleocapsid protein (red), and receptor-binding domain (green).

The modeled half-life under the gamma-decay model and the gamma-plateau model were also very similar and both models showed a rapid decay of the N-relative to the RBD- and S-antibody. The half-life for the N-, RBD- and S-antibody was 60 days [95% CI 52-68], 102 days [95% CI 92-114] and 126 days [95% CI 112-146] respectively under the gamma decay model, while the half-lives under the gamma-plateau model were 60 days [95% CI 52-70], 110 days [95% CI 74-148], and 364 days [95% CI 212-997] respectively. The half-lives under the gamma-plateau model were widened as a consequence of being closer to the time to plateau where the half-lives are stretched by a flattening curve (Figure 4 d and e).

Under the gamma-plateau model, the S-antibody was characterized by a slow decay, with an eventual stabilized plateau at 1825 days [95% CI 250-3700] and none of the posterior probability distribution of the titers at the eventual plateau crossed the threshold for a negative test, whereas 75% of the posterior probability distribution for the N-antibody crossed the threshold for a negative test by 610 days.

### Surrogate Neutralization Assay

There was a sigmoidal relationship between raw antibody titers and percentage binding/ACE-2 receptor blocking for both the S- and RBD-antibodies. The sigmoidal relationship demonstrated that above a threshold spike antibody titer of 8586 [95% CI 8160-9095] there was a dramatic increase in percentage binding/ACE-2 receptor blocking. When titer and blocking data were log transformed the relationship was linear with a strong positive correlation coefficient R^2^=0.72 and R^2^=0.77 for the spike antibody and the RBD respectively. In order to visualize the binding activity of spike and RBD antibodies at the plateau we mapped the maximum of the second derivative (the point at which the change in percentage binding increased the most) of the sigmoid curve to the final titers at the plateau (**Figure 5**). The point of maximum slope increase for the sigmoid curve was at 5430.11 [4553.50-6427.50] for the spike protein, and 5733.670 [4867.225-6423.525] for the receptor-binding domain. Whilst the full range of the distribution of spike antibodies were predicted to remain detectable indefinitely at plateau under the gamma-plateau model, only a small proportion of individuals were predicted to have titers sufficient to enable measurable functional binding under our surrogate neutralization assay.

**Figure 5:**
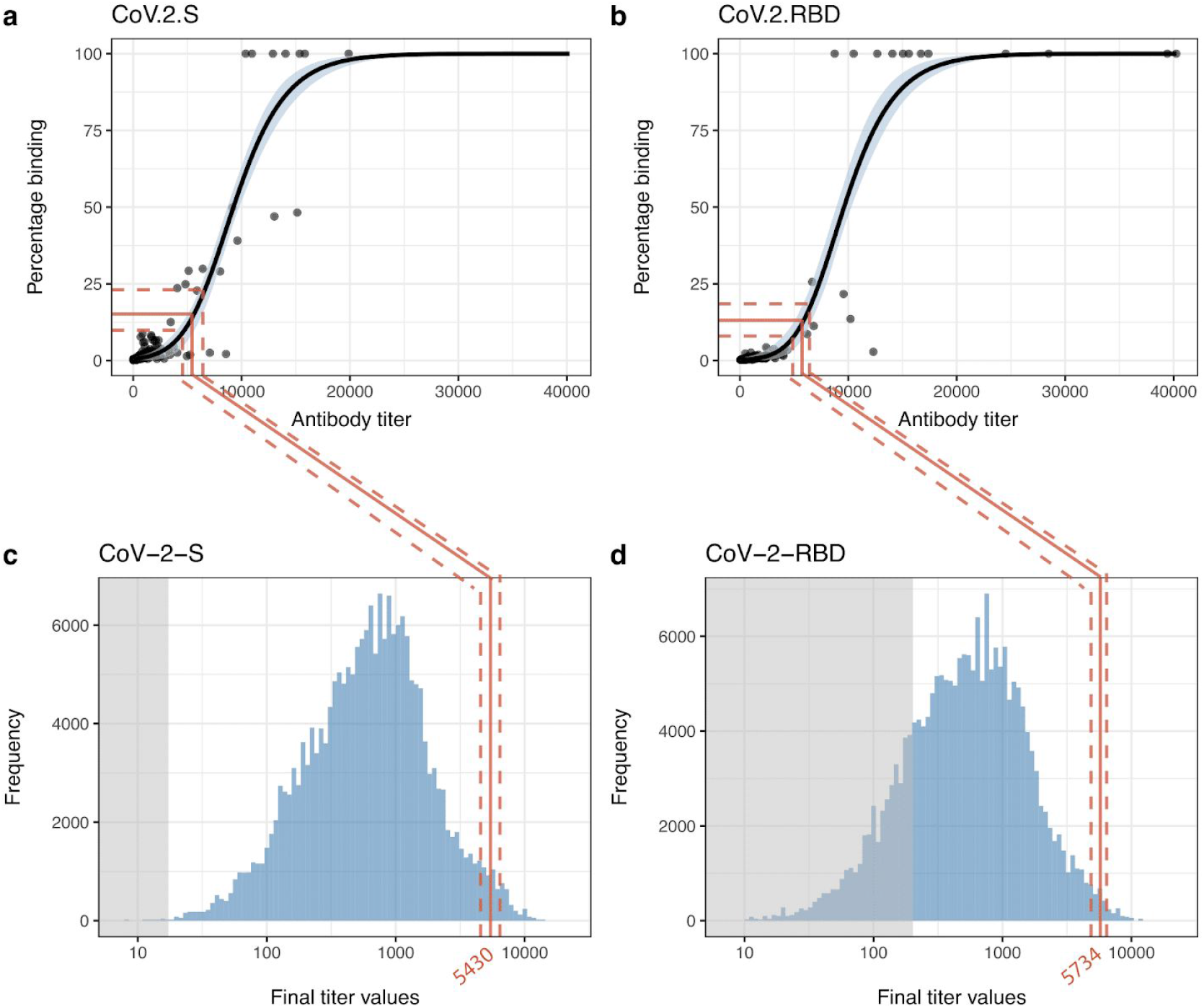
Surrogate neutralization assay (spike and RBD) and how this maps to the final predicted titers at plateau. **a,b** Percentage binding plotted against antibody titer. Red line represents the median amount of antibody titer at which the change in percentage binding is greatest, with the 95% CI indicated by dotted red lines. Black line is the median posterior distribution of the generalized logistic model, while the blue ribbon represents the 95% CI. **c,d**, Posterior distribution of the antibody titers at the long-term plateau as predicted by the gamma-plateau model. Shaded grey area corresponds to the threshold of seronegativity.

## Discussion

This prospective cohort study of antibody responses following SARS-CoV-2 infection has demonstrated that 99% of 349 healthcare workers symptomatic with SARS-CoV-2 remained seropositive for the spike protein antibody 200 days after symptoms. Our study is the first to provide a mathematical modeling framework capable of predicting the long-term dynamics of the 3 key SARS-CoV-2 antibodies following natural infection. Even under our most pessimistic assumptions of continuous exponential decay, 95% of individuals were predicted to remain seropositive to S-antibody at 465 days [95% CI 370-575 days] while our more optimistic upper bound gamma-decay model predicted a permanent long-lasting plateau of detectable S-antibody.

These data contradict conclusions from studies that have reported rapid waning of antibodies after a few months^29,31,32^. Furthermore, our findings are in line with the duration of humoral responses observed following SARS-CoV and MERS infections^17–20^. Importantly, the long-lasting S- and RBD-antibodies also correlated well with a surrogate SARS-CoV-2 neutralization assay of ACE-2-receptor-blocking, strongly suggesting that long-term measurable S-antibody levels are functionally important. When the humoral correlates of protection against reinfection are known, our model of longitudinal S-antibody dynamics will therefore enable predictions to be made about the duration of long-lasting protective immunity following infection with SARS-CoV-2.

In contrast to the S-antibody, the N-antibody was observed to serorevert in 56/349 participants over the course of the study alone and had a modeled half-life of 60 days. This has important implications for diagnostic testing, epidemiological modeling and public health decision making that often rely on the N-antibody to estimate SARS-CoV-2 seroprevalence. This finding may also explain some unexpectedly low population level prevalence estimates in high burden countries^37^ and confound the finding that children with multi-system inflammation have a higher S:N ratio compared to adults^30^. Antibodies peaked at 30-40 days, this is significantly longer than other reports that are likely to have missed the prolonged peak/plateau due to inadequate sampling density^38^. It is notable that the delay between the SARS-CoV-2 epidemic curve and the mini-epidemic of paediatric multi-system inflammatory syndrome coincides temporally with peak antibodies^39^.

The persistence of detectable S- and/or RBD-antibody compared to the rapid decay of the N-antibody has also been observed in convalescent sera obtained from SARS survivors, seventeen years after infection^19^, although the exact underlying mechanisms warrant further investigation. Differences in the epitope structure^40^, immunogenicity and presentation to B-cells may distinctly impact the production, maturation and longevity of the plasma cells that secrete these antibodies^41–44^. Distinct T-helper cell interactions at the germinal centre may further determine B-cell and humoral dynamics, as previously observed in the context of the response to different HIV proteins^45^. Independently or in parallel, transcriptional programs and epigenetic imprinting may also selectively influence the kinetics and survival of both N-versus S-antibody-producing long-lived plasma cells^43,44,46^. Finally, cross-reactive memory, rather than naive B cells, may play a role in responses targeting nucleoprotein, given that it is more conserved across CoVs than RBD^47^.

To date, no studies have comprehensively modeled the nature and duration of antibody responses to different SARS-CoV-2 epitopes. Long et al^26^, Seow at al^29^, Ibarrondo et al^27^, demonstrated rapid decay of SARS-CoV-2 IgG and neutralizing antibodies within the first 3 months following infection, particularly in mildly symptomatic cases. In July, our earlier publicly available pre-print of serial antibody and modeled data demonstrated longer lasting spike antibody and rapidly decaying N-antibody^48^. In comparison, others have reported that the S-antibody and/or RBD-antibody correlate with neutralizing responses and decay slowly, persisting during the study period, up to at 90-150 days post-infection^23–25,49^. These studies, however, are limited by their shorter sampling time frame, lower sampling density and lack of appropriate modeling to predict antibody trajectory. Implementing LOESS lines of best fit to the data^23–25,29^ or comparing the variance of average antibody titers at different time intervals^31,32,49^ does not permit evaluation of long-term antibody trajectory.

Our study is strengthened by the density, frequency, and duration of longitudinal sampling collection. The parallel evaluation of absolute antibody titers by the chemiluminescent MSD assay to three major SARS-CoV-2 proteins also enabled us to demonstrate the decay of the N-antibody relative to the S- and RBD-antibodies. Importantly, this is the first study to provide a mathematical framework for long-term SARS-CoV-2 antibody responses, modeling both the peak and decay following infection and enabling realistic best-case and worst-case predictions of future antibody titers. Our work provides a detailed, shareable and reproducible model, with parameters that are useful for epidemiological purposes. Additionally, some of our parameters are fitted at both the population and individual level which is informative when inspecting risk factors and variability in the population. A third possible trajectory may be that the humoral response stabilizes but then continues to decline (‘plateau then decay’), albeit at a slower rate, as previously demonstrated in the context of vaccine-induced HPV responses and Hepatitis A infection^35,36^. Further serological measurements of seropositive recruits will take place in 6 months’ time to confirm which of these three models is superior in the longer term.

None of the seropositive healthcare workers identified in this study required hospitalization. This is important, given that the overwhelming majority of COVID-19 cases are not hospitalized. Our study population is, therefore, representative of most community SARS-CoV-2 infections^50^. Severe disease has however been associated with higher antibody titers and a longer duration of antibody response following both SARS and MERS^17,21,51,52^.

To date, no definitive quantitative or qualitative correlate-of-protection has been identified for SARS-CoV-2 infection, disease or onward transmission. Nevertheless, findings from animal studies support the role of neutralizing antibodies as a correlate of anti-viral immunity^9,15,16^. Mapping our surrogate neutralization assay to the final distribution of antibody titers under the gamma-plateau model suggested that some measurable functional binding (surrogate neutralization) of antibody would occur at plateau albeit in a small proportion. Whether this level of long-term detectable antibody is sufficient to induce sterilizing immunity and limit transmission, or primarily attenuates severity of disease, remains to be seen. Formal neutralization assays >1-year post infection are required to clarify this further. Finally, SARS-CoV-2-specific T- and B-memory cellular responses must also be characterised to accurately determine durability of immunity. Indeed, robust memory T-cell responses to specific SARS-CoV-2 peptides are elicited following infection and their magnitude correlated with antibody titers^53–56^; nevertheless, these responses were also observed in seronegative and/or pauci-symptomatic individuals^53,56^. Prospective evaluation of re-infection alongside humoral, T-cell, B-cell and mucosal IgA/IgG/cellular dynamics should therefore be an urgent priority, particularly during the evolving second wave of the pandemic.

One potential limitation of this study is the fact that only 38% of participants had an available confirmatory positive PCR result. In order to mitigate this concern, prior to the study taking place, a formal performance evaluation of the MSD assay was undertaken among 169 confirmed PCR positive SARS-CoV-2 participants that demonstrated a 97.9% sensitivity and 97.4% specificity at 21 days post infection^57^. This makes the proportion of false positive serological tests likely to be small and therefore have little impact on our findings.

Viral neutralization assays remain the gold-standard *in vitro* correlate of protection; as such, the lack of formal ‘authentic’ neutralization tests is another limitation of the study. However, ACE-2 receptor competition assays, such as the MSD competitive binding assay, have been shown to correlate well with formal viral neutralization assays, enabling their use as a suitable surrogate functional test^58^.

Whilst the severity of infection among our study participants is likely to be representative of community infection, our findings may be biased to healthcare workers. Recent studies have hypothesized that previous exposure to seasonal CoVs - to which healthcare workers may be disproportionately exposed - may confer some protection against SARS-CoV-2^12,19,53–56,58^ and may need to be accounted for when modeling transmission or longevity dynamics^59^. Our estimates of the time-to-negativity are also dependent on the negative thresholds and lower limits of detection of the assay, respectively. However, our model fits, estimates of the rate of decay and the raw serial antibody titer trajectory (Figure 1) are not dependent on the threshold for a negative test.

In summary, this prospective cohort study has shown that the SARS-CoV-2 S-antibody, which correlated well with functional receptor blocking *in-vitro*, remained detectable in 99% of individuals up to 200 days post infection. In comparison, the N-antibody waned rapidly with a half-life of 60 days and 54/349 participants seroreverting over the course of the study. This study therefore has immediate consequences for diagnostic testing and public health decision making that often depend on the N-antibody as a reliable measure of past infection. Our most pessimistic continuous decay model, predicted that 95% of individuals would continue to have detectable spike antibody at 465 days while our gamma-plateau predicted that spike antibody would plateau at detectable levels indefinitely. The long-term presence of functional S (and RBD-antibody) has important implications for the duration of protective immunity following natural infection. It remains to be seen whether the SARS-CoV-2 vaccine candidates will replicate the long-lasting spike antibody duration observed and modeled here following natural infection.

## Materials and methods

### Study setting and design

Co-STARS is a 1 year single-centre, two-arm, prospective longitudinal cohort study of healthcare workers at a central London paediatric hospital Great Ormond Street Hospital for Children (GOSH). The study was approved to start by the United Kingdom NHS Health Research Authority on 29th April 2020 and registered on ClinicalTrials.gov (NCT04380896). Informed consent was obtained from all participants. The Study Protocol and Supplementary Materials submitted with this paper include detailed methods, power calculations and the data analysis approach.

### Study participants

All hospital staff members ≥18 years of age were eligible for the study, provided they did not display symptoms consistent with SARS-CoV-2 infection at recruitment. Those significantly immunosuppressed or those who had previously received blood products (including immunoglobulins or convalescent sera) since September 2019 were excluded from the study.

### Data Collection

After providing informed consent, participants undertook a detailed, standardised online questionnaire at study entry. This included socio-demographic factors, details of previous exposure to and symptomatic episodes consistent with COVID-19, any subsequent complications, previous SARS-CoV-2 diagnostic test results, past medical and contact history, and a comprehensive assessment of risk factors for exposure, susceptibility to infection and severe disease. Blood samples were also taken at baseline and each follow-up visit for determination of SARS-CoV-2 serology.

### Measurement of SARS-CoV-2 serum antibody and viral RNA by PCR

Serum antibodies titers were measured by the Meso Scale Discovery (MSD) Chemiluminescent binding assay that simultaneously detects and quantifies anti-SARS-CoV-2 IgG specific for trimeric S-protein, RBD and N-protein. Assay qualification and performance were evaluated as described in our accompanying methods paper^57^. Briefly, IgG levels for the MSD assay were expressed as arbitrary units calibrated against a set of reference sera distributed by the National Institute of Biological Standards and Control (Potters Bar, UK) under the auspices of the World Health Organisation. SARS-CoV-2 real-time reverse-transcriptase polymerase chain reaction (RT-PCR) targeting the N-gene was performed following RNA extraction, as previously described by colleagues at our laboratory^60^.

### Surrogate Neutralization Assay

An MSD® 96-well Custom Competition Assay designed to measure the inhibition of ACE-2 receptor binding to S or RBD by serum-derived antibody (MSD, Maryland) was run on 94 serial samples from 46 participants (two participants had 3 serial samples) in order to establish in vitro correlates of functional immunity.

### Follow-Up Appointments

All seropositive participants were followed up monthly (ongoing) for repeat antibody testing. Seronegative participants will be followed up 6-monthly. At each follow-up visit, participants completed a shortened version of the baseline questionnaire, focussing on any recurrent COVID-19 exposure and/or symptoms.

### Study outcomes

The primary outcome of the study was to establish humoral dynamics following SARS-CoV-2 infection. Nested sub-studies studies will explore the secondary outcome measures including the incidence of SARS-CoV-2 re-infection, the dynamics of the cellular response, IgA dynamics and the clinical and demographic factors that are associated with SARS-CoV-2 infection.

### Statistical Analysis and Power Calculations

Power calculations were based on a negative exponential model of antibody decay from the peak using the pwr.f2.test function in R. We assumed a study power of 80% and explored a variety of hypothesized effect sizes (decreases in antibody titers over 1 year) and co-variates on the required study size with an alpha of 0.05 (Supplementary Materials, Study Protocol).

### Statistical analysis and modeling of antibody dynamics

To calculate the proportion of individuals that serorevert over the course of the study, we performed a survival analysis to account for censoring using the survival package in R. An “event” was defined as a persistent negative test after the first positive test, while positive tests were counted as “censored” events.

The dynamics of antibody response following infection with SARS-CoV-2 were estimated by fitting two mixed effects models based on a gamma curve on every sample that had more than three antibody observations. Two gamma models were chosen (“gamma-plateau” and “gamma-decay”) to enable modeling of an optimistic upper bound estimate (eventual stabilization of decay to a plateau) to a pessimistic lower bound estimate (continuous exponential decay to zero). The gamma-decay model hypothesizes continuous antibody decay and does not account for long-lived humoral responses (although B-memory cells may still be induced to secrete antibody on challenge). On the other hand, the gamma-plateau model is based on the assumption that there are two phases of plasma cell production: ‘short-lived’ plasma cells are initially generated which secrete antibody with a short half-life, followed by a subsequent robust long-lived plasma cell (LLPC) response that maintains circulating, high-specificity, high-avidity antibody long-term^61^.

Gamma-decay and gamma-plateau models were fitted to the entire antibody response curve from day 0 (symptom onset) to the peak and then subsequent decay of each SARS-CoV-2 antibody. The confidence limits around the curves were derived by repeated sampling from the posterior distribution of the different model parameters, including individual effects.

The gamma-decay model assumed a uninterrupted continuous decay, given by the formula:

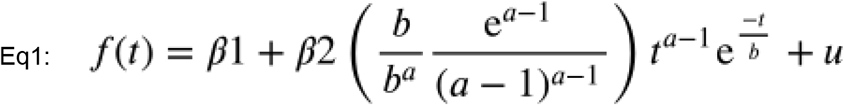

where f(t) is the log antibody titer at time t after symptom onset. The gamma function is described in terms of the shape (a) and scale (b), parameterized to reduce confounding of the parameters. β1 is the initial titer value at baseline; β2 determines the level of antibody rise; and u is an individual effect.

To account for the contribution of long lived plasma cells, a third term was added to the model, allowing for a long-term plateau expressed as:

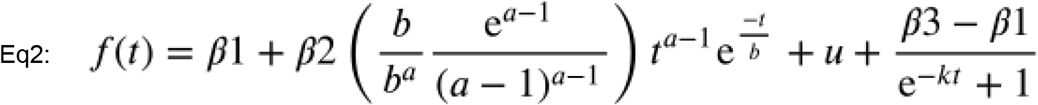

Where β3 represents the long-term plateau, and k is the rate at which the latter term rises.

The relationship between ACE-2 receptor blocking and antibody titers was modeled with a 4 parameter generalized logistic curve, where the percentage binding at titer level t is given by:

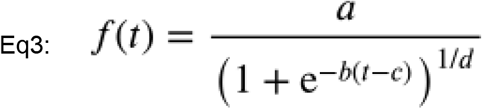

with parameters a,b, and c that represent, respectively, the upper receptor blocking asymptote, the growth rate, and the titer at which maximum growth occurs. The parameter d is an asymmetry factor that affects the point of inflection on the y axis.

All the models were fitted using RSTAN in R (R: A language and environment for statistical computing. Foundation for Statistical Computing, Vienna, Austria). For each model, we ran 4 independent chains for 15,000 iterations for the gamma distributions, and 10,000 iterations for the sigmoid model. Model comparison was performed using Pareto-smoothed importance sampling leave-one-out cross-validation (PSIS-LOO) as implemented in the loo R package.

### Ethics Statement

This study was approved by the UK Health Research Authority (www.hra.nhs.uk) and registered with www.clinical-trials.gov (NCT04380896). Written informed consent was obtained from all participants before recruitment to the study.

## Data Availability

Data available upon request.

## Acknowledgements

We would like to dedicate this article to the staff members who died of COVID-19 at Great Ormond Street Hospital during the first wave of the pandemic. We would also like to thank all the staff at Great Ormond Street Hospital who have taken part in the study. In addition, we are very grateful for all the hard work undertaken by the Great Ormond Street laboratory staff and the staff in the immunology laboratories both in the Camelia Botnar Laboratory and the Great Ormond Street Institute of Child Health who ensured that all the PCR tests and serological assays were completed in a timely manner. Finally, we would like to acknowledge the support of the Great Ormond Street Hospital Research & Development, Governance, Finance, Management, Estates, Operations and Communications departments.

## Supplementary data

**Supplementary Table S1.** Posterior distribution estimates and model convergence summary.

| Model | Antigen | Parameter | Mean | Median | 2.5% CI | 97.5% CI | ESS | R-hat |
| --- | --- | --- | --- | --- | --- | --- | --- | --- |
| Gamma-plateau | S | beta1 | 2.56 | 2.71 | 0.40 | 4.01 | 7290.13 | 1.00069 |
|  |  | beta2 | 0.93 | 0.85 | 0.33 | 1.96 | 19735.40 | 0.99997 |
|  |  | beta3 | 6.38 | 6.45 | 5.30 | 7.02 | 15862.35 | 1.00014 |
|  |  | a | 1.17 | 1.11 | 1.01 | 1.71 | 24185.03 | 1.00009 |
|  |  | b | 219.06 | 219.67 | 24.53 | 434.13 | 18160.67 | 1.00028 |
|  |  | k | 0.29 | 0.27 | 0.16 | 0.56 | 17365.52 | 1.00034 |
|  | RBD | beta1 | 2.35 | 2.40 | 0.37 | 4.14 | 7850.09 | 1.00015 |
|  |  | beta2 | 1.58 | 1.48 | 0.87 | 2.86 | 8768.42 | 1.00023 |
|  |  | beta3 | 6.18 | 6.34 | 4.81 | 6.82 | 8764.88 | 1.00026 |
|  |  | a | 1.20 | 1.11 | 1.00 | 1.92 | 16136.09 | 1.00003 |
|  |  | b | 135.27 | 112.05 | 34.75 | 344.81 | 9007.35 | 1.00018 |
|  |  | k | 0.29 | 0.27 | 0.17 | 0.53 | 17691.86 | 1.00001 |
|  | N | beta1 | 1.21 | 1.02 | 0.05 | 3.31 | 4898.48 | 1.00038 |
|  |  | beta2 | 3.78 | 3.73 | 2.56 | 5.23 | 3144.69 | 1.00087 |
|  |  | beta3 | 4.21 | 4.28 | 2.68 | 5.41 | 3289.29 | 1.00076 |
|  |  | a | 1.22 | 1.19 | 1.08 | 1.52 | 5808.56 | 1.00074 |
|  |  | b | 150.67 | 148.24 | 74.72 | 236.42 | 3335.84 | 1.00088 |
|  |  | k | 0.26 | 0.23 | 0.15 | 0.56 | 12731.13 | 1.00014 |
| Gamma-decay | S | beta1 | 1.27 | 0.93 | 0.03 | 4.37 | 4746.78 | 1.00094 |
|  |  | beta2 | 7.27 | 7.27 | 7.12 | 7.42 | 1142.77 | 1.00356 |
|  |  | a | 1.19 | 1.19 | 1.16 | 1.22 | 3218.21 | 1.00221 |
|  |  | b | 469.40 | 466.97 | 398.53 | 557.07 | 3230.69 | 1.00214 |
|  | RBD | beta1 | 0.99 | 0.71 | 0.03 | 3.46 | 4213.02 | 1.00126 |
|  |  | beta2 | 7.42 | 7.42 | 7.28 | 7.56 | 1042.43 | 1.00301 |
|  |  | a | 1.18 | 1.18 | 1.15 | 1.20 | 2729.89 | 1.00082 |
|  |  | b | 396.19 | 394.23 | 347.38 | 455.32 | 2735.57 | 1.00120 |
|  | N | beta1 | 1.01 | 0.73 | 0.03 | 3.51 | 4786.50 | 1.00096 |
|  |  | beta2 | 7.72 | 7.72 | 7.55 | 7.88 | 842.78 | 1.00572 |
|  |  | a | 1.21 | 1.21 | 1.19 | 1.24 | 3447.14 | 1.00217 |
|  |  | b | 253.60 | 253.15 | 232.89 | 276.65 | 3082.98 | 1.00252 |
| Sigmoid | S | a | 99.99 | 99.99 | 99.98 | 100.00 | 16995.30 | 1.00022 |
|  |  | b | 0.00 | 0.00 | 0.00 | 0.00 | 4278.59 | 1.00122 |
|  |  | **c** | 5854.63 | 5847.40 | 3829.62 | 7913.68 | 9890.67 | 1.00030 |
|  |  | **d** | 0.41 | 0.40 | 0.23 | 0.66 | 8331.01 | 1.00011 |
|  | **RBD** | **a** | 99.99 | 99.99 | 99.98 | 100.00 | 18844.79 | 1.00015 |
|  |  | **b** | 0.00 | 0.00 | 0.00 | 0.00 | 17737.25 | 1.00010 |
|  |  | **c** | 5286.62 | 5289.01 | 3354.71 | 7210.93 | 11869.92 | 1.00063 |
|  |  | **d** | 0.30 | 0.30 | 0.16 | 0.49 | 12695.78 | 1.00063 |

**Supplementary Table S2.** Model comparison summary.

| **Antigen** | **Model** | **WAIC** | **WAIC SE** | **ELPD** | **ELPD SE** |
| --- | --- | --- | --- | --- | --- |
| **S** | Gamma plateau | -1117.24 | 46.56 | -1129.73 | 47.57 |
| **S** | Gamma decay | -1170.36 | 39.18 | -1182.48 | 40.22 |
| **RBD** | Gamma plateau | -987.59 | 43.31 | -1000.95 | 44.61 |
| **RBD** | Gamma decay | -1061.868 | 38.38 | -1071.83 | 39.06 |
| **N** | Gamma plateau | -983.15 | 50.55 | -994.85 | 51.26 |
| **N** | Gamma decay | -1017.95 | 47.07 | -1030.21 | 48.19 |

**Supplementary Figure 1.**
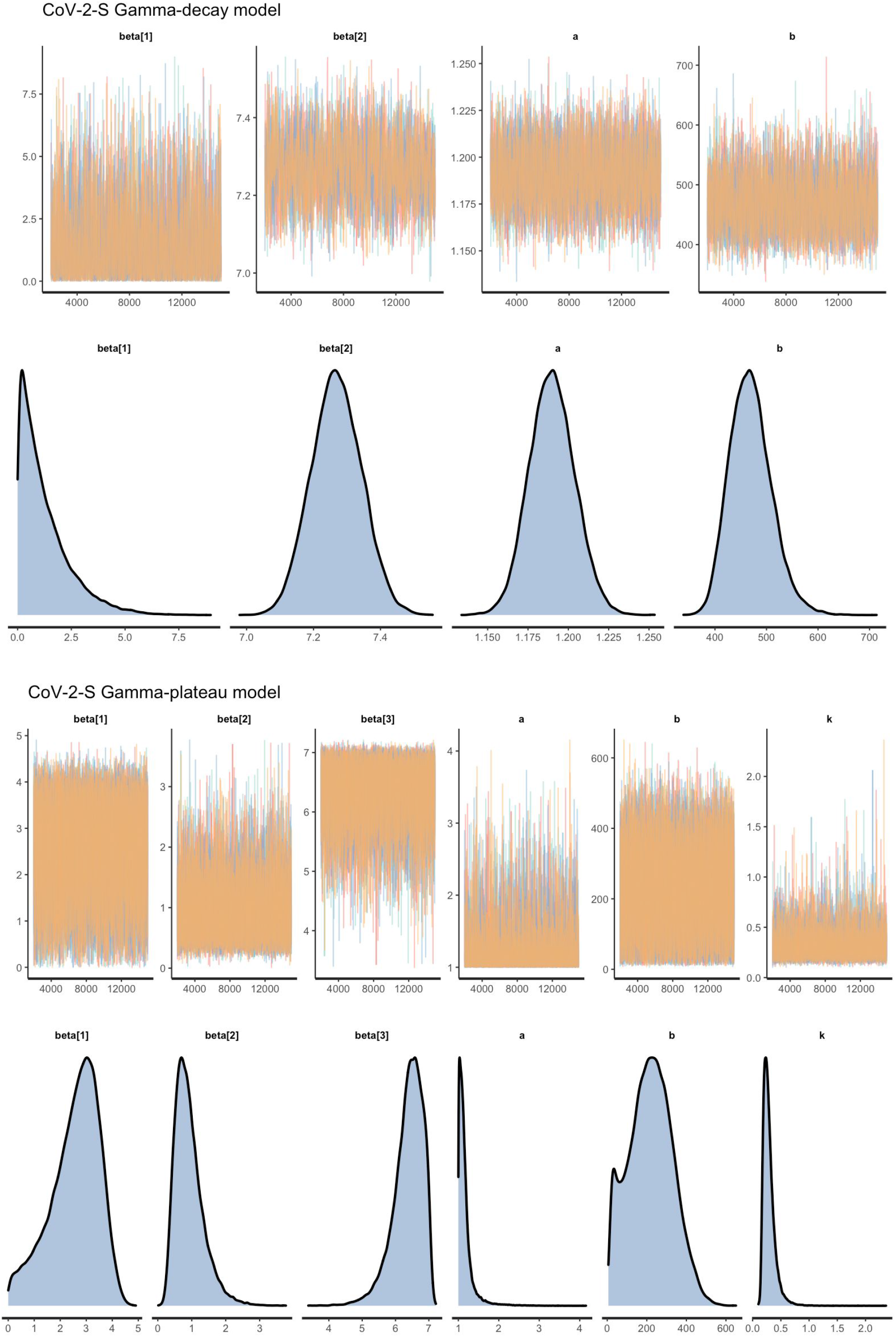
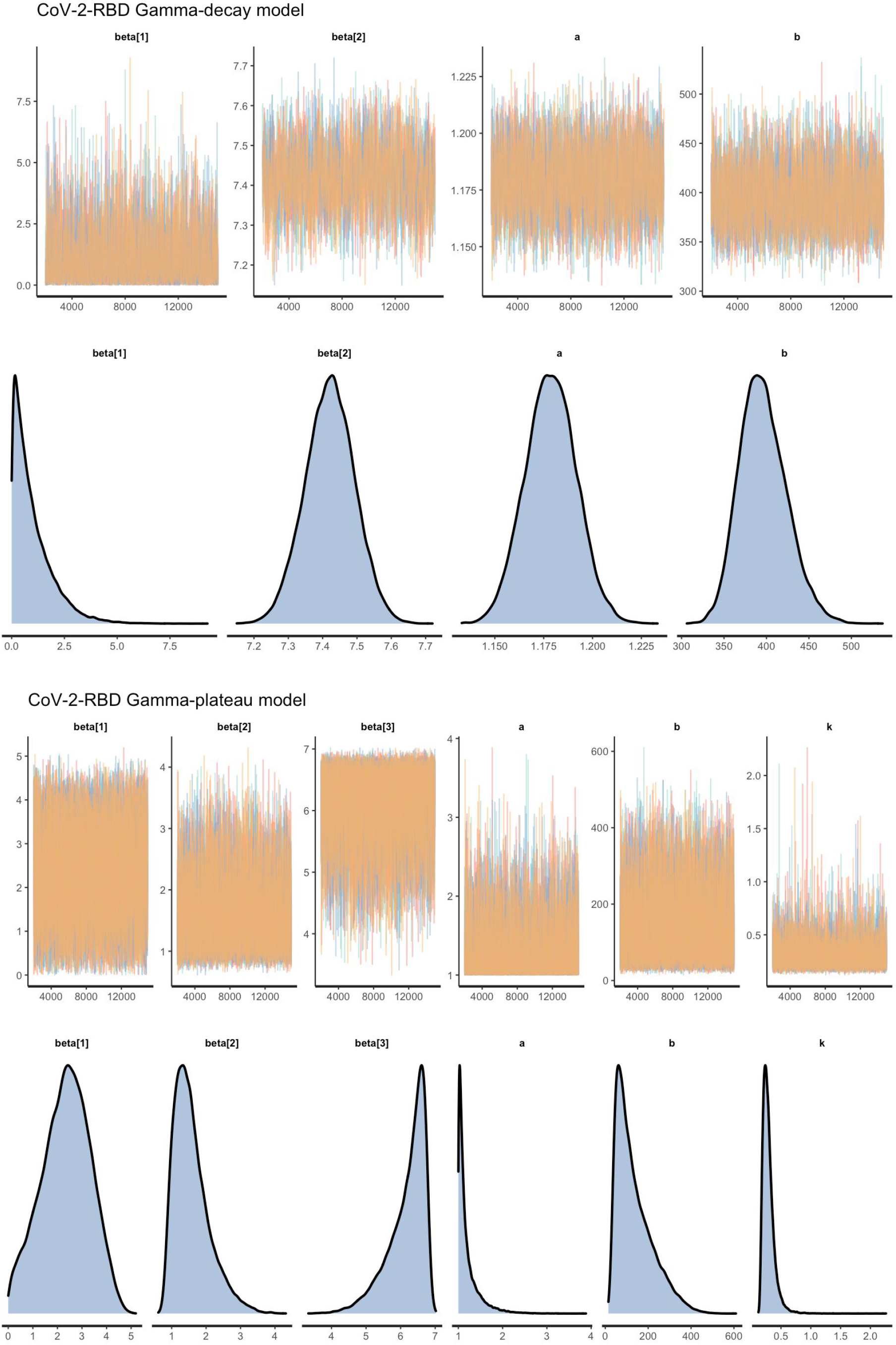
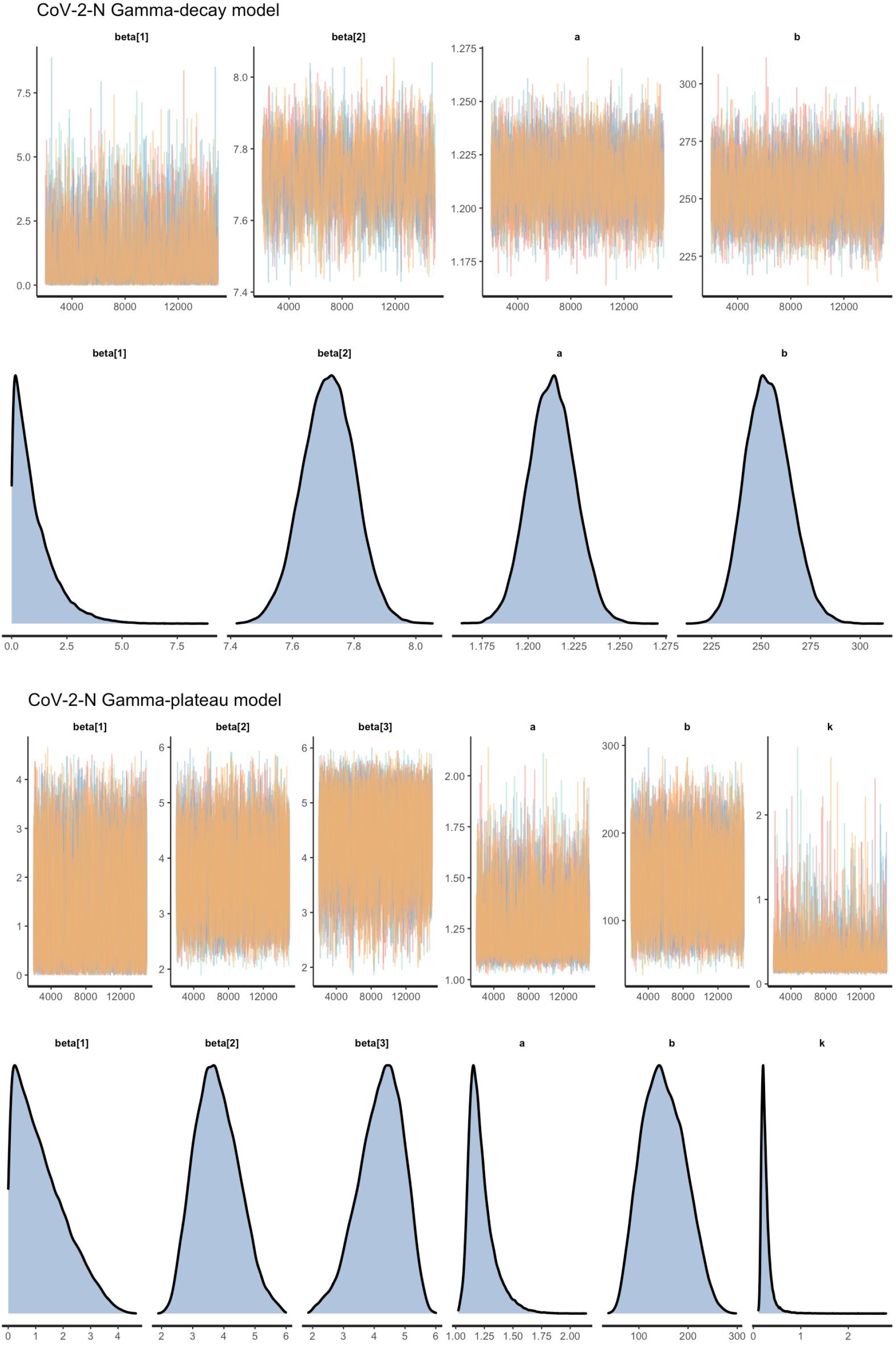
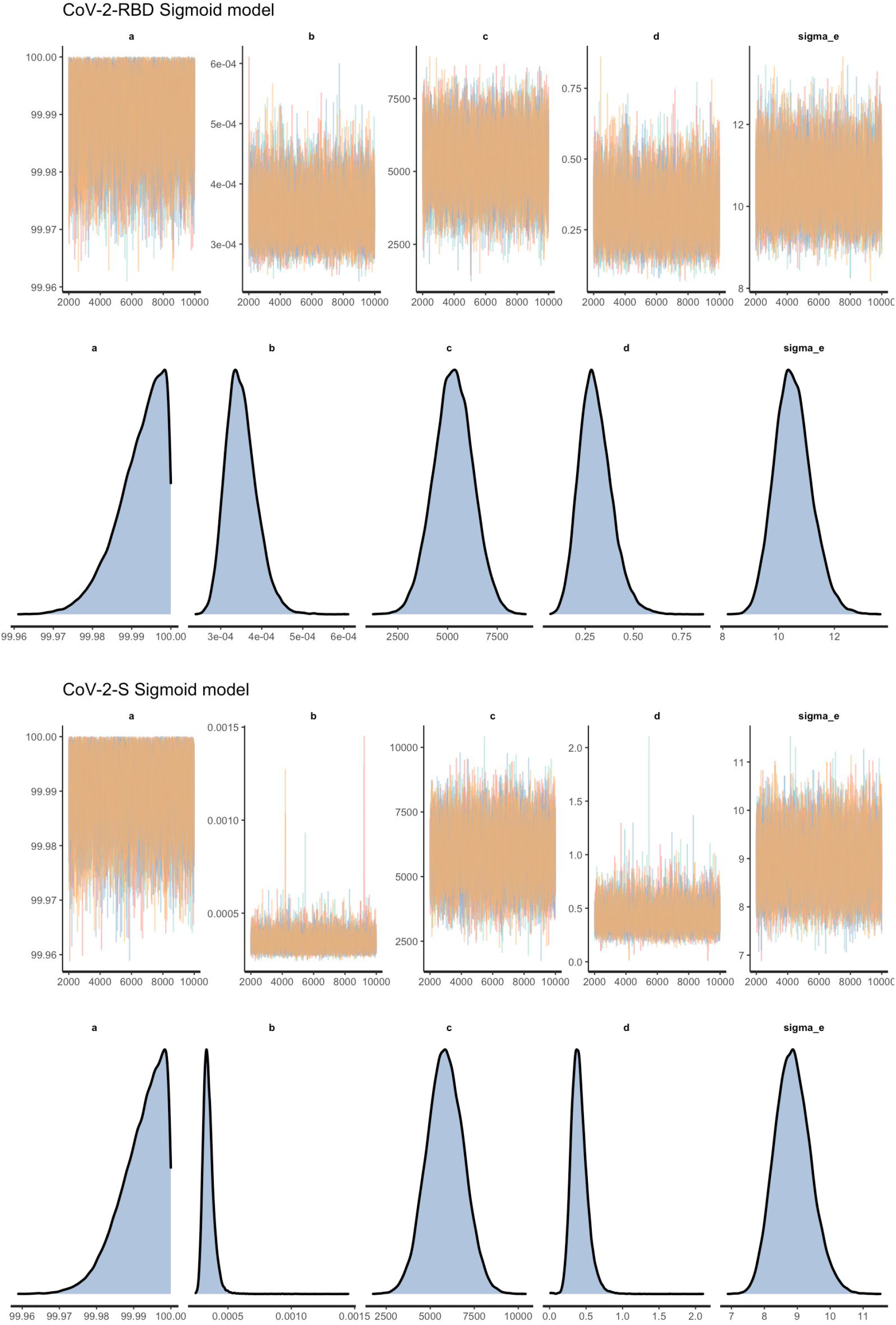
Monte Carlo Markov Chain Plots for Model Fits.

## References

1. Guan W, Ni Z, Hu Y, et al. Clinical Characteristics of Coronavirus Disease 2019 in China. New England Journal of Medicine [Internet] 2020 [cited 2020 Oct 30];382(18):1708–20. Available from:https://doi.org/10.1056/NEJMoa2002032

2. Dong E, Du H, Gardner L. An interactive web-based dashboard to track COVID-19 in real time. TheLancet Infectious Diseases [Internet] 2020 [cited 2020 Oct 30];20(5):533–4. Available from:https://www.thelancet.com/journals/laninf/article/PIIS1473-3099(20)30120-1/abstract

3. Zhu N, Zhang D, Wang W, et al. A Novel Coronavirus from Patients with Pneumonia in China, 2019. New England Journal of Medicine [Internet] 2020 [cited 2020 Oct 30];382(8):727–33. Available from:https://doi.org/10.1056/NEJMoa2001017

4. 12. Immune responses and immunity to SARS-CoV-2 [Internet]. European Centre for Disease Prevention and Control. [cited 2020 Oct 30];Available from: https://www.ecdc.europa.eu/en/covid-19/latest-evidence/immune-responses

5. Walls AC, Park Y-J, Tortorici MA, Wall A, McGuire AT, Veesler D. Structure, Function, and Antigenicity of the SARS-CoV-2 Spike Glycoprotein. Cell [Internet] 2020 [cited 2020 Oct 30];181(2):281-292.e6.Available from: https://www.ncbi.nlm.nih.gov/pmc/articles/PMC7102599/

6. Characterization of spike glycoprotein of SARS-CoV-2 on virus entry and its immune cross-reactivity with SARS-CoV | Nature Communications [Internet]. [cited 2020 Oct 30];Available from:https://www.nature.com/articles/s41467-020-15562-9

7. Cong Y, Ulasli M, Schepers H, et al. Nucleocapsid Protein Recruitment to Replication-Transcription Complexes Plays a Crucial Role in Coronaviral Life Cycle. Journal of Virology [Internet] 2020 [cited 2020Oct 30];94(4). Available from: https://jvi.asm.org/content/94/4/e01925-19

8. Ju B, Zhang Q, Ge J, et al. Human neutralizing antibodies elicited by SARS-CoV-2 infection. Nature[Internet] 2020 [cited 2020 Oct 30];584(7819):115–9. Available from:https://www.nature.com/articles/s41586-020-2380-z

9. Imai M, Iwatsuki-Horimoto K, Hatta M, et al. Syrian hamsters as a small animal model for SARS-CoV-2infection and countermeasure development. PNAS [Internet] 2020 [cited 2020 Oct30];117(28):16587–95. Available from: https://www.pnas.org/content/117/28/16587

10. Rogers TF, Zhao F, Huang D, et al. Isolation of potent SARS-CoV-2 neutralizing antibodies and protection from disease in a small animal model. Science [Internet] 2020 [cited 2020 Oct30];369(6506):956–63. Available from: https://science.sciencemag.org/content/369/6506/956

11. Suthar MS, Zimmerman MG, Kauffman RC, et al. Rapid Generation of Neutralizing Antibody Responses in COVID-19 Patients. Cell Reports Medicine [Internet] 2020 [cited 2020 Oct 30];1(3):100040. Availablefrom: https://linkinghub.elsevier.com/retrieve/pii/S2666379120300525

12. Ni L, Ye F, Cheng M-L, et al. Detection of SARS-CoV-2-Specific Humoral and Cellular Immunity inCOVID-19 Convalescent Individuals. Immunity [Internet] 2020 [cited 2020 Oct 30];52(6):971-977.e3.Available from: http://www.sciencedirect.com/science/article/pii/S1074761320301813

13. Gidari A, Nofri M, Saccarelli L, et al. Is recurrence possible in coronavirus disease 2019 (COVID-19)?. Case series and systematic review of literature. Eur J Clin Microbiol Infect Dis [Internet] 2020 [cited 2020 Oct 30];1–12. Available from: https://www.ncbi.nlm.nih.gov/pmc/articles/PMC7547550/

14. Tillett RL, Sevinsky JR, Hartley PD, et al. Genomic evidence for reinfection with SARS-CoV-2: a casestudy. The Lancet Infectious Diseases [Internet] 2020 [cited 2020 Oct 13];0(0). Available from:https://www.thelancet.com/journals/laninf/article/PIIS1473-3099(20)30764-7/abstract

15. Deng W, Bao L, Liu J, et al. Primary exposure to SARS-CoV-2 protects against reinfection in rhesus macaques. Science [Internet] 2020 [cited 2020 Oct 30];369(6505):818–23. Available from:https://science.sciencemag.org/content/369/6505/818

16. Chandrashekar A, Liu J, Martinot AJ, et al. SARS-CoV-2 infection protects against rechallenge in rhesus macaques. Science [Internet] 2020 [cited 2020 Oct 30];369(6505):812–7. Available from:https://science.sciencemag.org/content/369/6505/812

17. Cao W-C, Liu W, Zhang P-H, Zhang F, Richardus JH. Disappearance of antibodies to SARS-associated coronavirus after recovery. N Engl J Med 2007;357(11):1162–3.

18. Mo H, Zeng G, Ren X, et al. Longitudinal profile of antibodies against SARS-coronavirus in SARS patients and their clinical significance. Respirology 2006;11(1):49–53.

19. Chia WN, Tan CW, Foo R, et al. Serological differentiation between COVID-19 and SARS infections.Emerging Microbes & Infections [Internet] 2020 [cited 2020 Oct 30];9(1):1497–505. Available from:https://doi.org/10.1080/22221751.2020.1780951

20. Guo X, Guo Z, Duan C, et al. Long-Term Persistence of IgG Antibodies in SARS-CoV Infected Healthcare Workers. medRxiv [Internet] 2020 [cited 2020 Nov 3];2020.02.12.20021386. Available from:https://www.medrxiv.org/content/10.1101/2020.02.12.20021386v1

21. Payne DC, Iblan I, Rha B, et al. Persistence of Antibodies against Middle East Respiratory SyndromeCoronavirus. Emerg Infect Dis [Internet] 2016 [cited 2020 Oct 30];22(10):1824–6. Available from:https://www.ncbi.nlm.nih.gov/pmc/articles/PMC5038413/

22. Alshukairi AN, Khalid I, Ahmed WA, et al. Antibody Response and Disease Severity in Healthcare Worker MERS Survivors. Emerg Infect Dis 2016;22(6).

23. Isho B, Abe KT, Zuo M, et al. Persistence of serum and saliva antibody responses to SARS-CoV-2 spike antigens in COVID-19 patients. Science Immunology [Internet] 2020 [cited 2020 Oct 12];5(52). Availablefrom: https://immunology.sciencemag.org/content/5/52/eabe5511

24. Iyer AS, Jones FK, Nodoushani A, et al. Persistence and decay of human antibody responses to thereceptor binding domain of SARS-CoV-2 spike protein in COVID-19 patients. Science Immunology[Internet] 2020 [cited 2020 Oct 12];5(52). Available from:https://immunology.sciencemag.org/content/5/52/eabe0367

25. Ripperger TJ, Uhrlaub JL, Watanabe M, et al. Orthogonal SARS-CoV-2 Serological Assays Enable Surveillance of Low-Prevalence Communities and Reveal Durable Humoral Immunity. Immunity[Internet] 2020 [cited 2020 Nov 5];S1074761320304453. Available from:https://linkinghub.elsevier.com/retrieve/pii/S1074761320304453

26. Long Q-X, Tang X-J, Shi Q-L, et al. Clinical and immunological assessment of asymptomatic SARS-CoV-2 infections. Nature Medicine [Internet] 2020 [cited 2020 Oct 30];26(8):1200–4. Available from: https://www.nature.com/articles/s41591-020-0965-6

27. Ibarrondo FJ, Fulcher JA, Goodman-Meza D, et al. Rapid Decay of Anti–SARS-CoV-2 Antibodies in Persons with Mild Covid-19. New England Journal of Medicine [Internet] 2020 [cited 2020 Oct12];383(11):1085–7. Available from: https://doi.org/10.1056/NEJMc2025179

28. Loss of Anti–SARS-CoV-2 Antibodies in Mild Covid-19. New England Journal of Medicine [Internet]2020 [cited 2020 Nov 3];383(17):1694–8. Available from: https://doi.org/10.1056/NEJMc2027051

29. Seow J, Graham C, Merrick B, et al. Longitudinal evaluation and decline of antibody responses inSARS-CoV-2 infection. medRxiv [Internet] 2020 [cited 2020 Oct 12];2020.07.09.20148429. Availablefrom: https://www.medrxiv.org/content/10.1101/2020.07.09.20148429v1

30. Wajnberg A, Amanat F, Firpo A, et al. SARS-CoV-2 infection induces robust, neutralizing antibody responses that are stable for at least three months. medRxiv [Internet] 2020 [cited 2020 Oct13];2020.07.14.20151126. Available from:https://www.medrxiv.org/content/10.1101/2020.07.14.20151126v1

31. Crawford KHD, Dingens AS, Eguia R, et al. Dynamics of neutralizing antibody titers in the months after SARS-CoV-2 infection. J Infect Dis [Internet] [cited 2020 Nov 6];Available from:https://academic.oup.com/jid/advance-article/doi/10.1093/infdis/jiaa618/5916372

32. Tan Y, Liu F, Xu X, et al. Durability of neutralizing antibodies and T-cell response post SARS-CoV-2infection. Front Med [Internet] 2020 [cited 2020 Nov 3];Available from:https://doi.org/10.1007/s11684-020-0822-5

33. Wu J, Liang B, Chen C, et al. SARS-CoV-2 infection induces sustained humoral immune responses in convalescent patients following symptomatic COVID-19. medRxiv [Internet] 2020 [cited 2020 Oct13];2020.07.21.20159178. Available from:https://www.medrxiv.org/content/10.1101/2020.07.21.20159178v1

34. Zhao X, Ning Y, Chen MI-C, Cook AR. Individual and Population Trajectories of Influenza Antibody Titers Over Multiple Seasons in a Tropical Country. Am J Epidemiol [Internet] 2018 [cited 2020 Nov2];187(1):135–43. Available from: https://academic.oup.com/aje/article/187/1/135/3896092

35. Fraser C, Tomassini JE, Xi L, et al. Modeling the long-term antibody response of a human papillomavirus (HPV) virus-like particle (VLP) type 16 prophylactic vaccine. Vaccine [Internet] 2007[cited 2020 Nov 2];25(21):4324–33. Available from:http://www.sciencedirect.com/science/article/pii/S0264410X07002320

36. Andraud M, Lejeune O, Musoro JZ, Ogunjimi B, Beutels P, Hens N. Living on Three Time Scales: The Dynamics of Plasma Cell and Antibody Populations Illustrated for Hepatitis A Virus. PLOS Computational Biology [Internet] 2012 [cited 2020 Nov 2];8(3):e1002418. Available from:https://journals.plos.org/ploscompbiol/article?id=10.1371/journal.pcbi.1002418

37. Pollán M, Pérez-Gómez B, Pastor-Barriuso R, et al. Prevalence of SARS-CoV-2 in Spain (ENE-COVID):a nationwide, population-based seroepidemiological study. The Lancet [Internet] 2020 [cited 2020 Jul8];S0140673620314835. Available from:https://linkinghub.elsevier.com/retrieve/pii/S0140673620314835

38. Borremans B, Gamble A, Prager K, et al. Quantifying antibody kinetics and RNA detection during early-phase SARS-CoV-2 infection by time since symptom onset. eLife [Internet] 2020 [cited 2020 Nov 10];9:e60122. Available from: https://doi.org/10.7554/eLife.60122

39. 47. Multisystem Inflammatory Syndrome in Children in the United States. New England Journal of Medicine[Internet] 2020 [cited 2020 Nov 10];383(18):1793–6. Available from:https://doi.org/10.1056/NEJMc2026136

40. Slifka MK, Amanna IJ. Role of Multivalency and Antigenic Threshold in Generating Protective Antibody Responses. Front Immunol [Internet] 2019 [cited 2020 Nov 10];10. Available from:https://www.frontiersin.org/articles/10.3389/fimmu.2019.00956/full

41. Dörner T, Radbruch A. Antibodies and B Cell Memory in Viral Immunity. Immunity [Internet] 2007 [cited2020 Nov 10];27(3):384–92. Available from: https://www.cell.com/immunity/abstract/S1074-7613(07)00420-7

42. Amanna IJ, Slifka MK. Mechanisms that determine plasma cell lifespan and the duration of humoral immunity. Immunol Rev [Internet] 2010 [cited 2020 Nov 10];236(1):125–38. Available from:https://www.ncbi.nlm.nih.gov/pmc/articles/PMC7165522/

43. Khodadadi L, Cheng Q, Radbruch A, Hiepe F. The Maintenance of Memory Plasma Cells. FrontImmunol [Internet] 2019 [cited 2020 Nov 10];10. Available from:https://www.ncbi.nlm.nih.gov/pmc/articles/PMC6464033/

44. Nguyen DC, Joyner CJ, Sanz I, Lee FE-H. Factors Affecting Early Antibody Secreting Cell Maturation Into Long-Lived Plasma Cells. Front Immunol [Internet] 2019 [cited 2020 Nov 10];10. Available from:https://www.frontiersin.org/articles/10.3389/fimmu.2019.02138/full

45. Bonsignori M, Moody MA, Parks RJ, et al. HIV-1 Envelope Induces Memory B Cell Responses That Correlate with Plasma Antibody Levels after Envelope gp120 Protein Vaccination or HIV-1 Infection. TheJournal of Immunology [Internet] 2009 [cited 2020 Nov 10];183(4):2708–17. Available from:https://www.jimmunol.org/content/183/4/2708

46. Slifka MK, Ahmed R. Long-lived plasma cells: a mechanism for maintaining persistent antibody production. Curr Opin Immunol 1998;10(3):252–8.

47. Srinivasan S, Cui H, Gao Z, et al. Structural Genomics of SARS-CoV-2 Indicates Evolutionary Conserved Functional Regions of Viral Proteins. Viruses [Internet] 2020 [cited 2020 Nov 10];12(4):360.Available from: https://www.mdpi.com/1999-4915/12/4/360

48. Grandjean L, Saso A, Ortiz A, et al. Humoral Response Dynamics Following Infection with SARS-CoV-2.medRxiv [Internet] 2020 [cited 2020 Nov 10];2020.07.16.20155663. Available from:https://www.medrxiv.org/content/10.1101/2020.07.16.20155663v2

49. Wajnberg A, Amanat F, Firpo A, et al. Robust neutralizing antibodies to SARS-CoV-2 infection persist for months. Science [Internet] 2020 [cited 2020 Oct 30];Available from:https://science.sciencemag.org/content/early/2020/10/27/science.abd7728

50. Verity R, Okell LC, Dorigatti I, et al. Estimates of the severity of coronavirus disease 2019: a model-based analysis. The Lancet Infectious diseases 2020;0(0). - Google Search [Internet]. [cited 2020 Oct 31];Available from: https://www.google.com/search?q=Verity+R%2C+Okell+LC%2C+Dorigatti+I%2C+et+al.+Estimates+of+the+severity+of+coronavirus+disease+2019%3A+a+model-based+analysis.+The+Lancet+Infectious+diseases+2020%3B0(0).&rlz=1C5CHFA_enGB738GB738&oq=Verity+R%2C+Okell+LC%2C+Dorigatti+I%2C+et+al.+Estimates+of+the+severity+of+coronavirus+disease+2019%3A+a+model-based+analysis.+The+Lancet+Infectious+diseases+2020%3B0(0).&aqs=chrome..69i57.335j0j4&sourceid=chrome&ie=UTF-8

51. Ho M-S, Chen W-J, Chen H-Y, et al. Neutralizing Antibody Response and SARS Severity. Emerg InfectDis [Internet] 2005 [cited 2020 Oct 31];11(11):1730–7. Available from:https://www.ncbi.nlm.nih.gov/pmc/articles/PMC3367364/

52. Huang AT, Garcia-Carreras B, Hitchings MDT, et al. A systematic review of antibody mediated immunity to coronaviruses: kinetics, correlates of protection, and association with severity. NatureCommunications [Internet] 2020 [cited 2020 Nov 5];11(1):4704. Available from:https://www.nature.com/articles/s41467-020-18450-4

53. Le Bert N, Tan AT, Kunasegaran K, et al. SARS-CoV-2-specific T cell immunity in cases of COVID-19and SARS, and uninfected controls. Nature [Internet] 2020 [cited 2020 Nov 5];584(7821):457–62.Available from: https://www.nature.com/articles/s41586-020-2550-z

54. Juno JA, Tan H-X, Lee WS, et al. Humoral and circulating follicular helper T cell responses in recovered patients with COVID-19. Nature Medicine [Internet] 2020 [cited 2020 Nov 5];26(9):1428–34. Availablefrom: https://www.nature.com/articles/s41591-020-0995-0

55. Peng Y, Mentzer AJ, Liu G, et al. Broad and strong memory CD4 + and CD8 + T cells induced bySARS-CoV-2 in UK convalescent individuals following COVID-19. Nature Immunology [Internet] 2020[cited 2020 Nov 5];21(11):1336–45. Available from: https://www.nature.com/articles/s41590-020-0782-6

56. Grifoni A, Weiskopf D, Ramirez SI, et al. Targets of T Cell Responses to SARS-CoV-2 Coronavirus in Humans with COVID-19 Disease and Unexposed Individuals. Cell 2020;181(7):1489-1501.e15.

57. Johnson M, Wagstaffe HR, Gilmour KC, et al. Evaluation of a novel multiplexed assay for determining IgG levels and functional activity to SARS-CoV-2. J Clin Virol [Internet] 2020 [cited 2020 Oct14];130:104572. Available from: https://www.ncbi.nlm.nih.gov/pmc/articles/PMC7396134/

58. Wang C, Li W, Drabek D, et al. A human monoclonal antibody blocking SARS-CoV-2 infection. NatureCommunications [Internet] 2020 [cited 2020 Oct 31];11(1):2251. Available from:https://www.nature.com/articles/s41467-020-16256-y

59. Kissler SM, Tedijanto C, Goldstein E, Grad YH, Lipsitch M. Projecting the transmission dynamics ofSARS-CoV-2 through the postpandemic period. Science [Internet] 2020 [cited 2020 Oct31];368(6493):860–8. Available from: https://science.sciencemag.org/content/368/6493/860

60. Brown JR, Atkinson L, Shah D, Harris K. Validation of an extraction-free RT-PCR protocol for detection of SARS-CoV2 RNA. medRxiv [Internet] 2020 [cited 2020 Oct 31];2020.04.29.20085910. Available from:https://www.medrxiv.org/content/10.1101/2020.04.29.20085910v1

61. Cyster JG, Allen CDC. B Cell Responses: Cell Interaction Dynamics and Decisions. Cell2019;177(3):524–40.

